# Immunological Drivers and Potential Novel Drug Targets for Major Psychiatric, Neurodevelopmental, and Neurodegenerative Conditions

**DOI:** 10.1101/2024.02.16.24302885

**Authors:** Christina Dardani, Jamie W. Robinson, Hannah J. Jones, Dheeraj Rai, Evie Stergiakouli, Jakob Grove, Renee Gardner, Andrew M. McIntosh, Alexandra Havdahl, Gibran Hemani, George Davey Smith, Tom G. Richardson, Tom R. Gaunt, Golam M. Khandaker

## Abstract

Immune dysfunction is implicated in the aetiology of psychiatric, neurodevelopmental, and neurodegenerative conditions, but the issue of causality remains unclear, impeding attempts to develop new interventions. Using genomic data on protein and gene expression across blood and brain, we assessed evidence of a potential causal role for 736 immune response-related biomarkers on 7 neuropsychiatric conditions by applying Mendelian randomization (MR) and genetic colocalisation analyses. A systematic three-tier approach, grouping biomarkers based on increasingly stringent criteria, was used to appraise evidence of causality (passing MR sensitivity analyses, colocalisation, False Discovery Rate and Bonferroni thresholds). We provide evidence for a potential causal role of 29 biomarkers for 7 conditions. The identified biomarkers suggest a role of both brain specific and systemic immune response in the aetiology of schizophrenia, Alzheimers disease, depression, and bipolar disorder. Of the identified biomarkers, 20 appeared to be therapeutically tractable, including *ACE*, *TNFRSF17*, *SERPING1*, *AGER* and *CD40,* with drugs approved or in advanced clinical trials. Based on the largest available selection of plasma immune-response related biomarkers, our study provides insight into possible influential biomarkers for the aetiology of neuropsychiatric conditions. These genetically prioritised biomarkers now require further examination to evaluate causality, their role in the aetiological mechanisms underlying the conditions and therapeutic potential.

## Introduction

Psychiatric, neurodevelopmental, and neurodegenerative conditions (henceforth, neuropsychiatric conditions) are among the leading causes of disability worldwide^1,2^. These conditions are typically chronic, and affect mood, perception, cognition, and behaviour. Biological pathways contributing to these conditions are poorly understood, impeding attempts to identify effective new interventions^3^. For example, approximately one in three individuals with depression or schizophrenia do not respond to current medications which primarily target monoamine neurotransmitters^4^. This suggests that the current one-size-fits- all approach to treatment for these conditions may not be tenable. Therefore, identifying biological pathways underpinning neuropsychiatric conditions to help prioritise novel intervention targets remains a key priority for mental health research^5^.

Over the last two decades immune dysfunction has emerged as a promising mechanistic candidate for several neuropsychiatric conditions. For example, immune activating drugs induce depressive symptoms in hepatitis C patients^6^ and healthy volunteers^7^. Meta-analyses of case-control studies confirm atypical levels of cytokines in blood plasma and cerebrospinal fluid of individuals with schizophrenia, depression, and bipolar disorder^8,9^. Neuroimaging with positron emission tomography shows evidence of neuroinflammation in acute depression^10^. Nationwide cohort studies indicate associations between autoimmune conditions, infections and neuropsychiatric conditions, such as schizophrenia^11,12^, attention deficit hyperactivity disorder (ADHD)^13^, Alzheimer’s disease^14^, and depression^15^.

However, inferring causality remains an important outstanding issue because the observed associations between inflammation/immunity and neuropsychiatric conditions could be result of residual confounding or reverse causation. Mendelian randomisation (MR), a genetic causal inference method that can minimise these limitations by using genetic variants regulating levels/activity of biomarkers as proxies^16–18^, has provided some evidence for a potential causal effect of IL-6 and CRP in the onset of depression and schizophrenia^19,20^. RCTs suggest that broad spectrum anti-inflammatory drugs improve mood and psychotic symptoms in people with depression^21^ and schizophrenia^22^, but recent RCTs of monoclonal antibodies targeting specific cytokine pathways have yielded null findings^23–25^. This highlights the need for strengthening causal inference using complementary techniques and data sources to inform appropriate selection of therapeutic target/agent in future trials.

Furthermore, as existing studies have typically focused on a small number of immunological biomarkers, a comprehensive approach allowing investigations across hundreds of available biomarkers is necessary to obtain a more complete understanding of the role of immune dysfunction in neuropsychiatric conditions.

We examined evidence of a potentially causal role for 736 genetically proxied immunological biomarkers (i.e., all immune-response related biomarkers assayed in the plasma proteome) on the onset of seven major neuropsychiatric conditions (schizophrenia, bipolar disorder, depression, anxiety, ADHD, autism, and Alzheimer’s disease), using cutting-edge genomic causal inference methods, MR and genetic colocalisation. We harnessed quantitative trait loci (QTL) data, capturing protein abundance (pQTL) and protein-coding gene expression (eQTL) in blood and brain, to gain mechanistic insights into potential systemic (blood) and brain-specific effects. We conducted a series of sensitivity analyses to examine the robustness of our findings, including the possibility of reverse causation. We used a systematic three-tier approach to appraise evidence of causality, by grouping biomarkers based on increasingly stringent criteria. Evidence of causality was complemented by the assessment of therapeutic tractability of identified causal biomarkers to inform future translation.

## Materials & Methods

An overview of the analytic pipeline of the study can be found in Figure 1.

**Figure 1.**
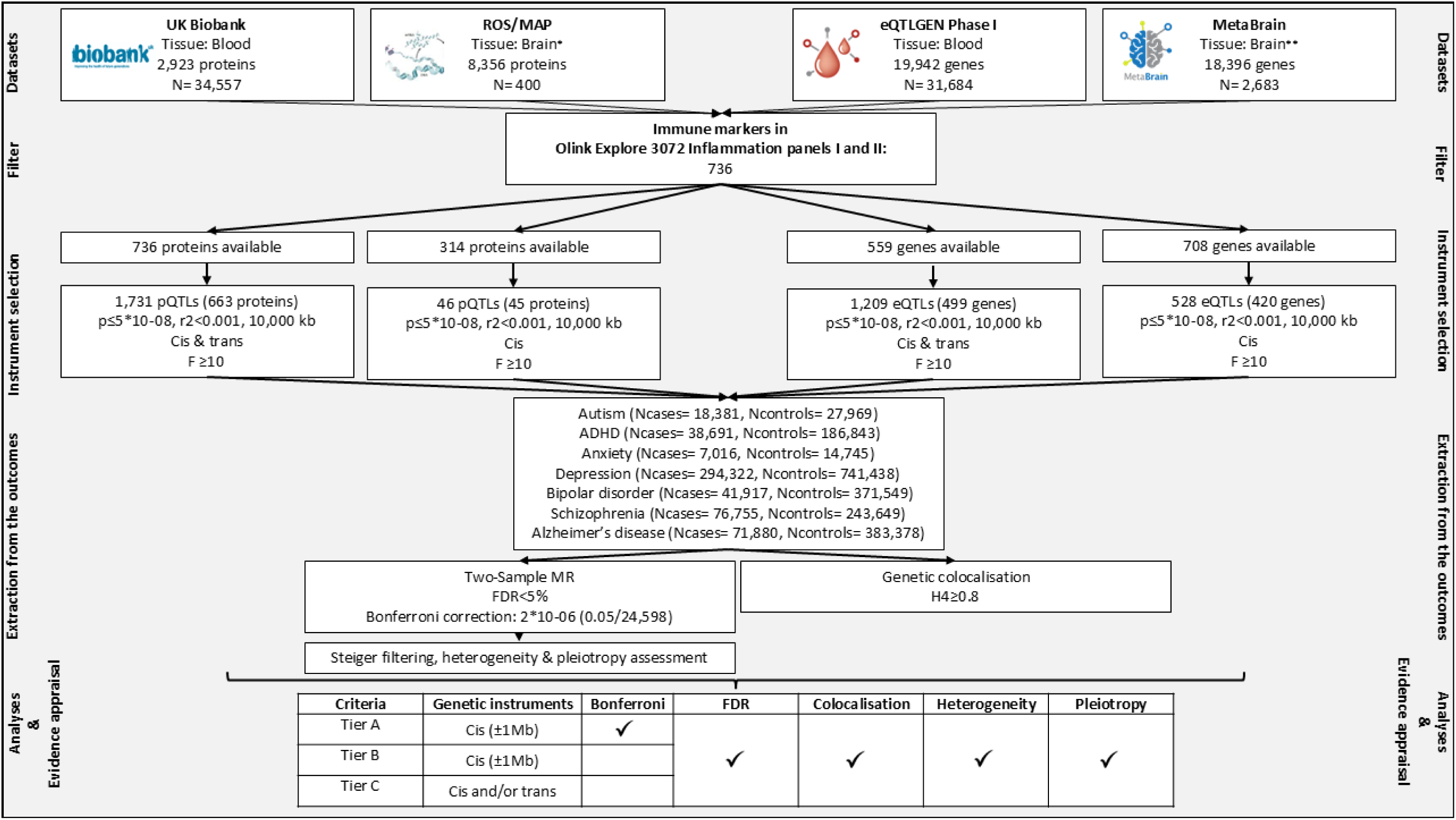
Analytic pipeline for assessing potentially causal immunological biomarkers for neuropsychiatric conditions and approach for evidence appraisal Note: *Dorsolateral prefrontal cortex (DLPFC), **Cortex.

### Genome-wide association studies (GWAS) of immunological biomarkers

#### Blood plasma derived protein abundance (blood pQTLs)

We used data from the largest genomic investigation of the human plasma proteome conducted in 34,557 European ancestry participants (discovery sample) of the UK Biobank (UKB)^26^ cohort, comprising 2,941 GWAS for 2,923 unique proteins assayed using the Olink Explore 3072 platform. Of these, we selected all proteins included in the Olink Inflammation panels I & II (n=736), which represent the most comprehensive collection of immunological biomarkers currently available (Supplementary Table 1). These 736 proteins formed the basis of our subsequent data extraction strategy from GWAS on blood and brain protein coding gene expression, and brain protein abundance. Details on the Olink Explore panels, assaying and genotyping in UKB can be found in the original publication^26^.

### Blood cell derived protein coding gene expression (blood eQTLs)

We used data from the eQTLGEN Phase I^27^ study of blood-cell derived gene expression in a sample of 31,684 individuals. The study assessed the expression of 19,942 genes, including 559 immunological protein coding genes present in the Olink Inflammation panels I & II.

### Brain tissue derived protein abundance (brain pQTLs)

To gain insights into potential brain specific effects, we additionally looked up the proteins of interest in the most comprehensive investigation of the brain proteome currently available^28^. It includes levels of 8,356 proteins in the dorsolateral prefrontal cortex (DLPFC) of 400 individuals from the Religious Orders Study (ROS) and Memory and Ageing Projects (MAP). Brain pQTLs were available for 314 immunological proteins present in the Olink Inflammation panels I & II.

### Brain tissue derived protein coding gene expression (brain eQTLs)

We used the latest meta-analysis of brain cortex gene expression (MetaBrain^29^) conducted in 6,601 RNA-seq samples (N=2,683) and covering 18,396 genes. This includes GWAS summary data for the expression of 708 genes encoding 708 proteins of interest in the brain cortex. Although MetaBrain provided data across different brain regions, the sample sizes were relatively small, and for this reason we used data for brain cortex rather than other brain regions (e.g., hippocampus, N= 168).

Further details on the study samples, genotyping and QTL analyses from the datasets used in the present study, can be found in the original publications^26,27,29,30^.

### GWAS of neuropsychiatric conditions

We used the latest (at the time of analysis) available GWAS summary data on seven neuropsychiatric conditions. Specifically, two neurodevelopmental: autism^31^ (Ncases= 18,381, Ncontrols= 27,969), attention deficit hyperactivity disorder (ADHD)^32^ (Ncases= 38,691, Ncontrols= 186,843); four psychiatric: anxiety^33^ (Ncases= 7,016, Ncontrols= 14,745), depression^34^ (Ncases= 294,322, Ncontrols= 741,438), bipolar disorder^35^ (Ncases= 41,917, Ncontrols= 371,549), schizophrenia^36^ (Ncases= 76,755, Ncontrols= 243,649); and one neurodegenerative condition: Alzheimer’s disease^37^ (Ncases= 71,880, Ncontrols= 383,378). Considering that the depression, bipolar disorder and Alzheimer’s disease GWASs included data from UKB, and this overlap with the GWASs of the plasma proteome may influence our findings, we additionally used available data on these three phenotypes excluding UKB for sensitivity analyses (depression: Ncases=166,773, Ncontrols= 507,679; bipolar disorder: Ncases= 40,463, Ncontrols= 313,436; Alzheimer’s disease: Ncases= 39,918, Ncontrols= 358,140). Details on the study samples, phenotype definition and genotyping can be found in the original publications^31–37^.

### Two-sample Mendelian randomization (MR)

MR utilises the special properties of germline genetic variants to strengthen causal inference within observational data^16^. Here we implemented MR as an instrumental variables analysis using common genetic variants as instruments. The method can yield unbiased causal effect estimates under assumptions that the instruments should satisfy: (1) they must be associated with the exposure, (2) they must not be associated with any confounders of the exposure outcome associations, (3) they should operate on the outcome entirely through the exposure (i.e., no horizontal pleiotropy)^38^.

For the present study, we performed two-sample MR, in which instrument-exposure and instrument-outcome effect sizes and standard errors were extracted from separate GWAS conducted in independent samples but representative of the same underlying population^39^.

### Instrument selection

For each exposure (immunological protein abundance, protein-coding gene expression), we used as genetic instruments common genetic variants that met the genome-wide significance threshold (p≤5*10^-08^) and were independent (r^2^<0.001; 10,000 kb). We assessed the strength of each instrument by estimating their F-statistic (F≥10 indicates adequate instrument strength)^40^. Genetic instruments with an F-statistic <10 were excluded to minimise weak instrument bias. As instruments, we selected a total of 1,731 plasma pQTLs for 663 unique proteins; 1,209 blood cell-derived eQTLs for 499 protein coding genes; 46 DLPFC pQTLs for 45 proteins; 528 brain cortex eQTLs for 420 protein-coding genes.

For blood pQTLs (UKB) and eQTLs (eQTLGEN), both *cis* and *trans* genetic instruments were available. Genetic instruments were categorised as *cis*-acting when they were located within proximity (±1Mb) to the gene regulatory region, and as *trans*-acting when located outside this window. Common genetic variants acting in *cis* to the protein-encoding gene are more likely to influence mRNA expression and protein levels (thus being less pleiotropic)^41^. On the other hand, *trans*-acting variants, are more likely to be pleiotropic due to their distance from the protein-encoding gene, but their inclusion can potentially increase the proportion of variance explained in the exposure, increasing the statistical power for MR analyses^41,42^. Acknowledging the above, we followed two complementary approaches: a biologically informed approach in which we used cis only instruments for our analyses, and a conventional approach allowing the inclusion of cis and/or trans instruments.

For brain pQTLs and eQTLs only *cis*-acting variants were used, because the Wingo et al.^28^, study reported *cis*-pQTLs only. The MetaBrain study reported only the statistically significant *trans*-eQTLs, without information on the respective regions around them, rendering genetic colocalization analyses for the *trans*-acting variants impossible (details on genetic instruments used across analyses in Supplementary Tables 2-5).

### Statistical Analyses

For each exposure, genetic instrument effect sizes and standard errors were extracted from each neuropsychiatric condition GWAS, and the variant-exposure, variant-outcome alleles were harmonised to ensure that effect sizes correspond to the same allele. If the exposure had only one associated variant, Wald ratio was used to generate causal effect estimates, and two-term Taylor expansion was used to approximate standard errors^29,43^. When more than one variant were available for an exposure, inverse variance weighted (IVW)^44^ regression was used.. In the case of IVW estimates we additionally assessed evidence on the heterogeneity of the genetic instruments using the Cochran’s Q statistic as well as the potential pleiotropy influencing the causal effect estimates using the Egger intercept. Details on the Wald ratio and IVW methods as well as Cochran’s Q and Egger intercept can be found in Supplementary Note 1. We used the Benjamini-Hochberg method^45^ to control false discovery rate (FDR) across our analyses (FDR<5%). In addition, we appraised findings using a strict Bonferroni-corrected p-value threshold 2*10^-06^ (0.05/ 24,598).

### Genetic colocalisation

Colocalisation analysis can complement MR by elucidating a distinct aspect of the identified causal relationship between an exposure and an outcome^46^. Specifically, colocalisation allows the assessment of the hypothesis that any identified causal effects are driven by the same causal variant influencing both exposure and outcome, instead of distinct causal variants that are in linkage disequilibrium (LD) with each other^47^. In practice, the approach harnesses SNP coverage within the same specified locus for two traits of interest and tests whether the association signals for each trait at the locus are suggestive of a shared causal variant^47^.

Considering that genetic instruments used in our analyses comprised either single variants (MR Wald ratio estimates) or multiple *cis* and/or *trans* variants (MR IVW estimates), we followed three distinct approaches for variant selection for colocalisation analyses to help prioritise (where possible) variants with the highest biological relevance for the exposure of interest. Specifically, when the instrument consisted of variants of which at least one was *cis*, the *cis* variant(s) was tested for colocalisation. If the instrument consisted of multiple *trans* variants, we used the *trans* variant with the smallest p-value for colocalisation. Finally, if the instrument was a single variant, the variant was tested for colocalisation regardless of whether it was *cis* or *trans*.

We extracted regions within ±500KB around the instrumented variant and implemented the algorithm described by Robinson *et al*^48^ to perform pairwise conditional and colocalisation (PWCoCo) analysis, which assesses all conditionally independent signals in the exposure dataset region against all conditionally independent signals in the outcome data. Genotype data from mothers in the Avon Longitudinal Study of Parents and Children (ALSPAC) cohort^49^ were used as the LD reference panel (N= 7,733; for ALSPAC cohort details and available genotype data see Supplementary Note 2). We ran these analyses using the default settings, as suggested by the authors in the original publications^42,47^. Evidence of colocalisation was considered if there was an H4 posterior probability of both traits having a shared causal variant ≥ 0.8, as proposed by the authors of the method. Although we did not exclude the wider Major Histocompatibility Complex (MHC) region for these analyses, we marked the genetic instruments that were found to reside within it (25–34DMb) and recommend caution when interpreting their colocalisation evidence due to the complex linkage disequilibrium (LD) structure of the region^50^.

### Steiger filtering

We performed Steiger filtering to assess whether causal effect estimates were influenced by reverse causation^51^. The method assesses whether the genetic variants proxying the exposure explain more variance in the outcome, which, if true, suggests that the primary phenotype influenced by the variant is the outcome rather than the exposure.

### Drug target prioritisation and validation

We used a three-tier system to prioritise evidence of causality for the biomarkers, and to explore their potential as drug targets. Tier A included findings that had evidence which passed the Bonferroni threshold (≤1.8*10^-06^), passed Steiger filtering, and passed colocalisation (H4≥0.8). Tier B included findings that passed the FDR threshold (<%5), passed Steiger filtering, and passed colocalisation (H4≥0.8). Importantly, for both Tiers A and B, the biomarker had to be proxied by genetic instruments that were *cis* variants (i.e., only findings from the biologically informed approach). Tier C included findings where the MR analyses allowed the inclusion of *trans* variants and fulfilled the same requirements as Tier B. Across all tiers, it was additionally required for the genetic instruments (if n instruments >2) to show no evidence of heterogeneity as defined by the Cochran’s Q statistic and the MR estimates (if n instruments >3) no evidence of pleiotropy as defined by the Egger intercept.

When there was Tier A, B or C evidence for a biomarker across different QTLs (eQTL & pQTL) and tissue types (brain & blood), we performed genetic colocalisation analyses using PWCoCo between the QTLs of the biomarker. This approach allowed us to investigate whether the effects on the outcome were driven by the same underlying variant across QTLs and tissue types, which increases reliability of that molecular marker’s relationship with the condition^52^. These analyses were not conducted in cases that the QTLs were residing in the MHC region.

We looked up potential therapeutic tractability of the identified Tier A, B or C biomarkers using small molecule, antibody binding, and/or any other treatment modality^53^ in the Open Targets Platform (https://platform.opentargets.org/). The Open Targets Platform is a freely available online resource for drug target identification and prioritisation that integrates genetic and genomic data with existing evidence on protein structure and function, and information on approved drugs, and ongoing clinical trials^53,54^. Open Targets have categorised target tractability based on eight buckets/groups for small molecules and nine buckets/groups for antibodies (see https://github.com/chembl/tractability_pipeline_v2 ). In order to aid interpretation, we categorised tractability into three mutually exclusive groups, in line with previous work^55^: Group 1. Strong druggability evidence: buckets 1, 2 & 3 for small molecules, antibodies, other modalities; Group 2. Likely or potentially druggable: buckets 4-8 for small molecules, 4 & 5 for antibodies; Group 3: Little or unknown druggability: remaining buckets. Data retrieved on 21/01/2025.

### Enriched pathways and phenotypes for the identified causal biomarkers

To aid the interpretation of our findings and elucidate potential biological pathways underlying the identified causal biomarkers, we performed gene-set enrichment analyses using GeneNetwork^56^. Developed by the MetaBrain consortium, GeneNetwork allows enrichment analyses using terms available on the Gene Ontology (GO), KEGG and Reactome pathway resources, as well as the Human Phenotype Ontology (HPO) database. We used the GeneNetwork browser (analyses conducted on 21/01/2025) and entered in the analyses the biomarkers that satisfied the Tier A, B and C criteria for each neuropsychiatric condition. These analyses were conducted only for neuropsychiatric conditions with at least five identified causal biomarkers (satisfying Tier A, B, or C criteria).

### Bi-directional two-sample MR

To assess reverse causation, we tested the causal effects of genetic liability to each neuropsychiatric condition on circulating immunological proteins. Genetic instruments for each condition were extracted from the respective GWAS using a p-value threshold of ≤5*10^-08^ (r^2^<0.001; 10,000 kb). The only exceptions were autism and anxiety, for which a p-value threshold of ≤5*10^-07^ was used as there were insufficient instruments at the genome-wide significance threshold (2 and 1 instrument respectively). Details on the instruments for each neuropsychiatric condition can be found in Supplementary Table 6. Genetic instruments were then extracted from the GWAS of the immunological proteins (736 proteins)^26^ and their alleles were harmonised. Causal effects were estimated using the IVW approach (Supplementary Note 1)^44^. Due to the number of tests conducted (7 phenotypes*736 immunological biomarkers available in UKB) we used Bonferroni correction (p≤9.7*10^-06^).

### Software

Analyses were carried out using the computational facilities of the Advanced Computing Research Centre of the University of Bristol (http://www.bris.ac.uk/acrc/). Blood plasma pQTL data and blood cell derived eQTL data were extracted and processed using the gwasvcf package version 1.0 in R (https://github.com/MRCIEU/gwasvcf)^57^. The summary data from MetaBrain were lifted over from GRCh38 to GRCh37 using the UCSC liftover tool^58^ to match the build of the rest of the data. Two-sample MR, Steiger filtering, and bi-directional MR analyses were conducted using functions from the TwoSampleMR R package version 0.5.6 (https://github.com/MRCIEU/TwoSampleMR)^59^ and the mrpipeline R package (https://github.com/jwr-git/mrpipeline). The PWCoCo algorithm was implemented using the Pair-Wise Conditional analysis and Colocalisation analysis package v1.0 (https://github.com/jwr-git/pwcoco)^48^. Pathway and phenotype enrichment analyses were conducted using the GeneNetwork browser v2.0 (https://www.genenetwork.nl/)^56^.

## Data availability

Across all analyses published summary-level data were used and no patient identifiable information was included. UKB blood pQTL data can be accessed through the portal: http://ukb-ppp.gwas.eu. Brain pQTL data can be accessed through the Synapse portal: https://www.synapse.org/#!Synapse:syn24172458. Blood eQTL data can be accessed through https://www.eqtlgen.org/phase1.html. Brain cortex eQTL data can be accessed through the MetaBrain platform: https://www.metabrain.nl/. GWAS data on ADHD, autism, anxiety, bipolar disorder, and schizophrenia can be accessed at: https://pgc.unc.edu/for-researchers/download-results/. GWAS data on depression can be accessed at: https://ipsych.dk/en/research/downloads/. GWAS data on Alzheimer’s disease can be accessed at: https://ctg.cncr.nl/software/summary_statistics.

## Code availability

A version of the code at time of publication will be available at the study-dedicated GitHub repository. For the purposes of the manuscript review, please find the link: https://github.com/ChristinaDni/immunological_drivers_psychiatric_mr_pwcoco

## Results

### Immunological drivers for neuropsychiatric conditions

In total, we found evidence for 151 potentially causal relationships corresponding to 83 unique immunological biomarkers that passed the FDR<5% (Supplementary Table 7). Among these 29 unique biomarkers met our strict Tier A, B or C criteria for causal evidence (Figure 2). Eight biomarkers (4 unique proteins and 4 genes) met the most stringent Tier A criteria (i.e., passed the stringent Bonferroni threshold: 1.82*10^-06^, Steiger filtering, showed no evidence of heterogeneity and pleiotropy (in cases of IVW estimates), were supported by evidence of colocalisation, and consisted of *cis* variants). Seventeen biomarkers (5 unique proteins and 12 genes) met Tier B criteria (i.e., passed the FDR threshold, Steiger filtering, showed no evidence of heterogeneity and pleiotropy (in cases of IVW estimates), were supported by evidence of colocalisation, and the instruments consisted of *cis* variants). Seven effects corresponding to 6 unique proteins and one gene, met Tier C criteria (genetic instruments consisted of *cis* and/or *trans* variants and fulfilled the Tier B requirements).

**Figure 2.**
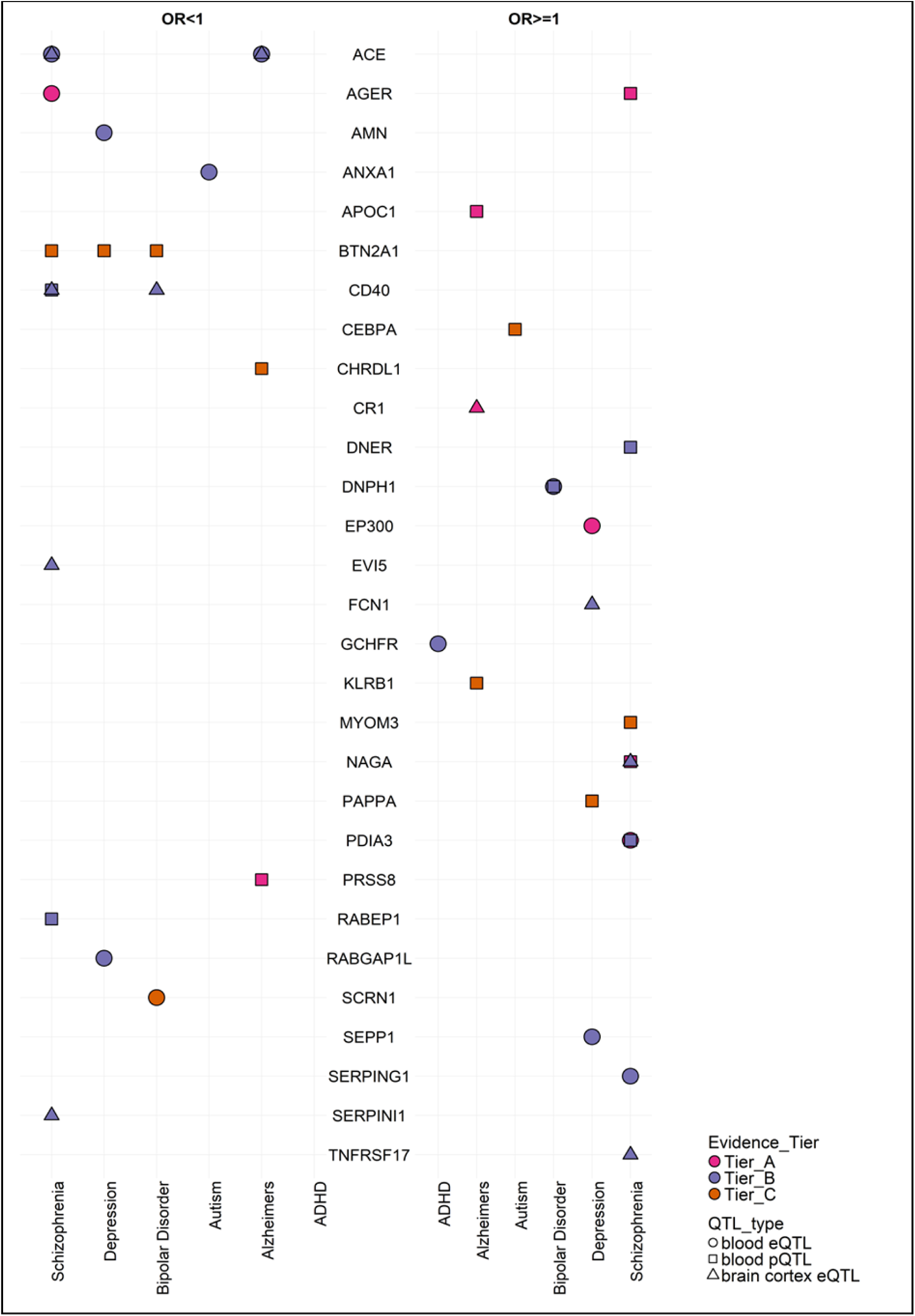
Summary of the Tier A, B and C findings of the analyses investigating potential effects of genetically proxied immunological biomarkers across neuropsychiatric conditions.

Schizophrenia (n=57) and Alzheimer’s disease (n=28), followed by depression (n=24), bipolar disorder (n=24) and had the highest number of potentially causal immunological markers after FDR correction.

### Condition specific findings

#### Neurodevelopmental conditions

For autism, we identified 6 potentially causal immunological biomarkers after FDR correction, two of which met Tier B/C criteria. The effect of genetically proxied expression of *ANXA1* in blood tissue (FDR= 0.01; H4>0.8) fulfilled the Tier B criteria. Genetically proxied levels of CEBPA in blood fulfilled the Tier C criteria (FDR=0.01, H4>0.8). For ADHD, 10 potentially causal immunological biomarkers were identified after FDR, of which genetically proxied expression of *GCHFR* in blood fulfilled the Tier B criteria (FDR=0.01; H4>0.8). Detailed MR and colocalisation findings are available in Supplementary Tables 7 and 8.

#### Psychotic and affective disorders

We found 57 potentially causal immunological biomarkers for schizophrenia after FDR correction, of which 4 met the Tier A criteria. Specifically, we found evidence for genetically proxied expression of *PDIA3* in the blood (p= 7*10^-08^, H4>0.8) along with genetically proxied levels of NAGA in the blood (p=6.9*10^-10^, H4>0.8). There was also evidence for genetically proxied expression and levels of *AGER* in the blood (p=1.8*10^-09^, p= 6.9*10^-14^, H4>0.8, respectively), although these results should be treated with caution as the lead SNPs for *AGER* reside in the MHC region. In addition, 12 met the Tier B criteria. Specifically, we found evidence of potential causal effects for genetically proxied expression of *ACE* in blood (FDR=0.007; H4>0.8) and brain cortex (FDR=0.002; H4>0.8), genetically proxied expression of *SERPING1* (FDR= 0.002; H4>0.8) in blood, genetically proxied expression of *TNFRSF17* (FDR=0.02; H4>0.8), *CD40* (FDR=0.004; H4>0.8), *SERPINI1* (FDR=0.04; H4>0.8), *EVI5* (FDR=0.003, H4>0.8) and *NAGA* (FDR= 0.001, H4>0.8) in brain cortex, and genetically proxied levels of RABEP1 (FDR= 0.01; H4>0.8), DNER (FDR=0.005; H4>0.8), PDIA3 (FDR= 0.003, H4>0.4) and CD40 (FDR= 0.05, H4>0.8) in blood. Furthermore, BTN2A1 (FDR= 5*10^-15^; H4>0.8) and MYOM3 (FDR=0.01, H4>0.8) fulfilled the Tier C criteria, both based on blood *trans* pQTLs (Supplementary Tables 7 and 8).

Among the 24 potentially causal immunological biomarkers identified for depression, genetically proxied expression of *EP300* in blood satisfied the tier A criteria (p=2*10^-09^, H4>0.8). Moreover, genetically predicted expression of *AMN* (FDR=0.03, H4>0.8), *SEPP1* (FDR=0.04, H4>0.8), *RABGAP1L* (FDR= 0.004, H4>0.8) in blood and *FCN1* (FDR= 0.02, H4>0.8) in brain cortex, satisfied the tier B criteria. Genetically proxied levels of BTN2A1 (FDR= 4.8*10^-06^, H4>0.8) and PAPPA (FDR= 0.009, H4>0.8) in blood (FDR=4*10^-06^; H4>0.8) fulfilled the Tier C criteria. These findings were largely consistent in sensitivity analyses using the depression GWAS excluding UKB ( Supplementary Tables 7 and 8).

For bipolar disorder, we found 24 potentially causal biomarkers. Among these, genetically proxied expression of *SCRN1* (p=1.05*10^-06^, H4>0.8) in blood satisfied the tier A criteria, while genetically predicted expression of *CD40* (FDR=0.002; H4>0.8) in brain cortex satisfied the tier B criteria. Genetically proxied expression and levels of *DNPH1* in blood (FDR=0.04; FDR= 0.03, H4>0.8, respectively) fulfilled the Tier B criteria. Genetically proxied levels of BTN2A1 in blood (FDR=3.8*10^-05^, H4>0.8) fulfilled the Tier C criteria. In sensitivity analyses using the bipolar disorder GWAS excluding UKB, estimates were consistent and confidence intervals overlapping with our main findings (Supplementary Tables 7 and 8).

For anxiety, no estimated causal effects of genetically proxied immunological biomarkers surpassed the FDR threshold (<5%; Supplementary Tables 7 and 8).

### Alzheimer’s disease

We identified 28 potentially causal biomarkers for Alzheimer’s disease. Among these, genetically proxied expression of *CR1* (FDR= 5.3*10^-14^; H4>0.8) in the brain cortex fulfilled the Tier A criteria as well as genetically predicted levels of APOC1 (p=2.4*10^-96^, H4>0.8) and PRSS8 (p= 3.4*10^-07^, H4>0.8) in blood. In addition, genetically proxied expression of *ACE* in blood (FDR= 0.005; H4>0.8) and brain (FDR=0.001; H4>0.8) fulfilled the Tier B criteria.

Genetically proxied levels of KLRB1 (FDR=0.001, H4>0.8) andCHRDL1 (FDR= 2.5*10^-05^) fulfilled the Tier C criteria. In sensitivity analyses using the Alzheimer’s disease GWAS excluding UKB, the estimates for the effect of APOC1 were directionally consistent with our main findings but the confidence intervals were not overlapping (Supplementary Tables 7 and 8).

### Drug target identification, prioritisation, and validation

In total, 20 unique biomarkers meeting our strict Tier A, B or C criteria for causal evidence appeared to be therapeutically tractable (Supplementary Table 9). Notably, *ACE*, which had Tier B evidence for both schizophrenia and Alzheimer’s, has approved drugs for cardiovascular indications. *AGER* (Tier A evidence for schizophrenia) and *CD40* (Tier B evidence for both schizophrenia and bipolar disorder) have drugs in advanced clinical trials. Furthermore, *TNFRSF17* (Tier B evidence for schizophrenia) *SERPING1* (Tier B evidence for schizophrenia) have approved drugs. See Supplementary Table 10 for details on drugs approved or in clinical trials for *ACE*, *AGER*, *CD40, SERPING1,* and *TNFRSF17*.

Across our analyses, five immunological biomarkers were supported by evidence from different QTL types. Specifically, the effects of *ACE* on schizophrenia and Alzheimer’s disease were supported by brain cortex and blood eQTLs, the effects of *CD40* on schizophrenia were supported by blood pQTLs and brain cortex eQTLs, the effects of *NAGA* on schizophrenia were supported by blood pQTLs and brain cortex eQTLs, while the effects of *DNPH1* on bipolar disorder were supported by blood pQTLs and eQTLs. For each biomarker the direction of effect estimates across QTL types was convergent (Supplementary Figure 1), while there was evidence of colocalisation between them, suggesting that the identified effects are likely to be driven by the same underlying variant (Supplementary Table 11). It is worth noting here that although we had evidence from blood pQTLs and eQTLs for a potential effect of *AGER* on schizophrenia, we did not test for colocalisation between the QTLs due to the lead SNPs residing in the MHC region.

### Enriched pathways and phenotypes for the identified causal biomarkers

Analyses were conducted for schizophrenia, depression, and Alzheimer’s disease as these had enough (≥5) biomarkers satisfying the Tier A, B, or C criteria. The findings of the enrichment analyses can be found in Supplementary Table 12. For schizophrenia, 13 biomarkers were entered in GeneNetwork, and six for depression and Alzheimer’s disease (respectively). In the case of schizophrenia, there were some patterns in the enrichment findings related to reproductive phenotypes (e.g., HP: infertility, p= 2*10^-04^) and pathways implicated in microbial response (e.g., REACTOME: Beta defensins, p= 4*10^-05^). In the case of depression, there were some patterns in the findings related to fatty acid phenotypes (e.g., HP: Abnormal fatty acid concentration, p= 5*10^-04^) and pathways involved in NF-kappaB signalling (e.g., GO: negative regulation of I-kappaB kinase/NF-kappaB signalling, p= 6*10^-08^). In the case of Alzheimer’s disease enrichment findings were less consistent with the exception of some evidence of enrichment for some ageing-related phenotypes (e.g., ptosis, p= 7*10^-05^)..

### Evidence of reverse causation

In bidirectional MR analyses, none of the estimated causal effects of genetic liability to autism, ADHD, schizophrenia, bipolar disorder, or anxiety on levels of immunological proteins, surpassed the Bonferroni correction threshold (p≤9.7*10^-06^). However, we found evidence of causal effects of genetic liability to depression on levels of CXCL17 (p=1.7*10^-07^), and PRSS8 (p=6.6*10^-06^). Similarly, we found that genetic liability to Alzheimer’s disease had causal effects on levels of APOF (p=2.4*10^-07^), and IL32 (p=7*10^-11^). See Supplementary Table 13.

## Discussion

Recent decades have seen limited progress in new therapeutics for neuropsychiatric conditions. Despite converging evidence implicating immune dysfunction in several neuropsychiatric conditions, the success of immunotherapy clinical trials remains elusive. One key barrier is the lack of a clear understanding of causality to inform appropriate selection of therapeutic target/agent. In this study, using cutting-edge genomic causal inference methods applied to largescale proteomic and gene expression data from blood and brain, we have assessed evidence for potential causality for the largest available selection of immune-response related biomarkers in relation to the onset of seven neuropsychiatric conditions. We provide evidence for causality for 29 immunological biomarkers providing evidence suggesting that both brain specific and systemic immune response may contribute to pathogenesis of neuropsychiatric conditions, especially schizophrenia, Alzheimer’s disease, depression, and bipolar disorder.

Among the 29 identified immunological biomarkers, eight satisfied the strictest criteria for potential causality (Tier A). Specifically, *AGER*, *PDIA3* and NAGA appeared to have an effect on schizophrenia. Existing evidence suggests that the three genes are implicated in glycosylation^60–62^. Glycosylation is a complex biological process related to the production of glycans, and has been recently hypothesised to be implicated in the aetiology of schizophrenia^63^. In the case of Alzheimer’s disease, *CR1* and APOC1 appeared to have Tier A evidence of effects, in line with existing literature implicating them in the aetiopathogenesis of condition^64,65^. *SCRN1*, identified to have effects on bipolar disorder, is a novel phosphorylated tau binding protein that has been shown to be abundant in amyloid plaques^66^ and has been recently identified as shared in cross-trait analyses between bipolar disorder and inflammatory bowel disease^67^. Similarly, in depression, *EP300*, satisfying Tier A evidence, has been identified in cross-trait analyses as shared between depression and insomnia^68^.

### From prioritised biomarkers to drug targets for neuropsychiatric conditions

Among the biomarkers prioritised in the present project, we found that 20 of them are potentially druggable. Among them, *AGER* (schizophrenia), *CD40* (schizophrenia & bipolar), *TNFRSF17* (schizophrenia), *ACE* (schizophrenia & Alzheimer’s) and *SEPRING1* (schizophrenia) have drugs approved or in advanced clinical trials for several indications including cardiovascular and autoimmune conditions. Before deriving conclusions on the potential opportunities for drug repurposing, the present findings should be viewed in the context of important biological and methodological considerations outlined below.

### Pathways from transcription to translation

A small proportion of the identified biomarkers were linked to neuropsychiatric conditions via gene expression and protein abundance. Specifically, the effects of *DNPH1* on bipolar disorder were via gene expression and protein abundance in blood, the effects of *PDIA3* on schizophrenia were via gene expression and protein abundance in blood, while the effects of *NAGA* and *CD40* on schizophrenia were via protein abundance in blood and gene expression in brain cortex. In addition, the direction of the identified effects was concordant across gene expression and protein abundance which is encouraging when it comes to drug target validation and prioritisation^52^.

However, a large number of our findings were not supported by both protein abundance and gene expression and in cases that it did, the effect estimates were discordant (this was the case for the effects of AGER on schizophrenia via gene expression and protein abundance in blood). This can substantially impact the potential of the identified biomarkers as drug targets. One possible explanation for this are differences in power across the datasets (e.g., the brain QTL data were based on a sample of 400 individuals). Another possibility may be alternative splicing events. Alternative splicing has a central role in the pathway from transcription to translation as it results in the production of multiple proteins via different signalling pathways^69^. Alternative splicing events may play an important role in neuropsychiatric conditions, such as schizophrenia ^70^. Future investigations incorporating datasets that capture the pathway from transcription to translation (i.e, eQTLs, sQTLs and pQTLs) are necessary to further validate the potential of the current prioritised biomarkers as drug targets, particularly considering that most existing drugs act via protein activity rather than gene expression.

### Tissue specific effects

A number of the identified biomarkers had effects on neuropsychiatric conditions via QTLs measured in blood. This suggests that not only brain-specific immunological processes are important in these conditions, but also systemic^71^. In addition, two of the prioritised markers (*CD40* and *ACE*) were supported by effects of the biomarkers measured in blood as well as brain cortex. Although this might seem encouraging with regards to potential therapeutic applications, it is difficult to derive conclusions from the present evidence. Specifically, *CD40* has low tissue specificity and *ACE* is predominantly expressed in the small intestine. Drug targets from genes with low tissue specificity (*CD40* in this case) or genes that have enhanced expression in tissues other than the one investigated (*ACE* in this case) have the risk of leading to off-target side effects^72,73^. A careful investigation of the identified biomarkers in the context of their tissue-enhanced expression is necessary in order further understand their potential as drug targets.

### Effects across neuropsychiatric conditions

In the case of *ACE* and *CD40* we found evidence of effects on more than one neuropsychiatric condition. Specifically, we found that decreased expression of *ACE* in blood and brain cortex is linked to increased risk of both schizophrenia and Alzheimer’s disease.

This is consistent with results from previous MR studies^74,75^. Considering that ACE inhibitors are widely used for the management of hypertension, these findings require further investigation. The identified effect for Alzheimer’s particularly may be a result of survival bias, considering that hypertension can lead to early mortality and therefore individuals may not live long enough to be diagnosed with the condition^76,77^. Beyond its effects on hypertension, ACE inhibition in rats leads to memory and learning impairments^78^. Therefore, another possibility is that ACE inhibition does not causally influence risk to the conditions per se, but some of their common phenotypic expressions, such as cognitive decline, which is common to both schizophrenia and Alzheimer’s disease. Therefore, choosing the right outcome would be as important as choosing the right drug target in future RCTs. Similarly, *CD40* expression in brain may influence risk of both schizophrenia and bipolar disorder by causally influencing psychotic symptoms, which are common to both conditions. These possibilities require further investigation.

### Effects on developmental stage and progression

Our study design allowed us to investigate the causal effects of immunological biomarkers on the onset of neuropsychiatric conditions but not progression^79^. Identifying actionable treatment targets can be particularly complex for conditions with neurodevelopmental origins such as schizophrenia, where pathogenic changes could take place well before the emergence of clinical symptoms^80^. Therefore, the question of the potential utility of the identified targets in conditions with neurodevelopmental origins remains, and requires careful consideration of conceptual, methodological, and ethical aspects.

### Limitations

Our study has some methodological limitations. First, the study was conducted using GWAS data of European ancestry individuals, and therefore generalisability of our findings to other populations remains a concern. Second, although we used the largest GWAS data available the possibility of limited statistical power cannot be excluded, particularly for brain pQTL data, and the anxiety GWAS. Third, methodological discrepancies across datasets (e.g., mass spectrometry in brain pQTLs, antibody-based methods in blood pQTLs), might have influenced the potential of the study to identify converging effects across blood and brain.

Fourth, our enrichment analyses were based on a small number of biomarkers (13 in schizophrenia, 6in depression and Alzheimer’s), which may limit the reliability of these findings. Fifth, UKB proteomic GWAS had some sample overlap with some of the neuropsychiatric conditions, notably Alzheimer’s disease (>50%), depression (34%), and bipolar disorder (14%). Sample overlap can introduce a bias toward the observational estimate^81^, though this is unlikely to adversely influence hypothesis testing^82^. Moreover, in sensitivity analyses using Alzheimer’s, depression and bipolar disorder GWAS excluding UKB the effect estimates were consistent with the ones derived from the primary analyses and the confidence intervals were overlapping. Sixth, though our results are suggestive of causal relationships we are unable to prove causality due to potential horizontal pleiotropy or violations of other MR assumptions such as gene-environment equivalence and consistency of treatment effects. Further studies using different data sources (e.g., measured levels of the biomarkers) and methodological approaches aligning with the principles of causal triangulation^83^ in diverse populations (e.g., in terms of ancestry and/or age groups) are necessary in order to establish causality. Finally, although we assessed the possibility of reverse causation, it is difficult from these analyses to derive conclusions on the individual potentially causal relationships between genetic liability to neuropsychiatric conditions and levels of biomarkers. This can be a promising avenue for research focusing on understanding the health-related outcomes of these conditions and future work allowing in depth investigations (e.g., assessing the potential influence of pleiotropy across conditions) of these relationships is necessary.

## Conclusions

Using a comprehensive analytic approach allowing the integration of genomic data on protein and gene expression across blood and brain, we identify a potential causal role for 29 immunological biomarkers on seven neuropsychiatric conditions. However, considering the complexity of the phenotypes examined, further investigations of the identified biomarkers are required, using different data sources, methodological approaches and diverse populations to establish their potential role in the aetiology of neuropsychiatric conditions. This way we can expect new and better treatments for individuals with neuropsychiatric conditions.

## Data Availability

All GWAS data used in this work is publicly available for research purposes. Individual-level data from the ALSPAC birth cohort are not publicly available for reasons of clinical confidentiality. Data can be accessed after application to the ALSPAC Executive Team who will respond within 10 working days. Application instructions and data use agreements are available at http://www.bristol.ac.uk/alspac/researchers/access/.

## Ethics Statement

ALSPAC data used as reference panel: ethical approval was obtained from the ALSPAC Ethics and Law Committee and the Local Research Ethics Committees. We are extremely grateful to all the families who took part in this study, the midwives for their help in recruiting them, and the whole ALSPAC team, which includes interviewers, computer and laboratory technicians, clerical workers, research scientists, volunteers, managers, receptionists, and nurses. GWAS data: across all analyses summary data were used. These data are publicly available for research purposes. Details on the ethics declarations for each GWAS study can be found in the original publications.

